## Extended Data Figures for "ARF1 prevents aberrant type I IFN induction by regulating STING activation and recycling"

**Extended Data Fig. 1** **Hirschenberger et al.**

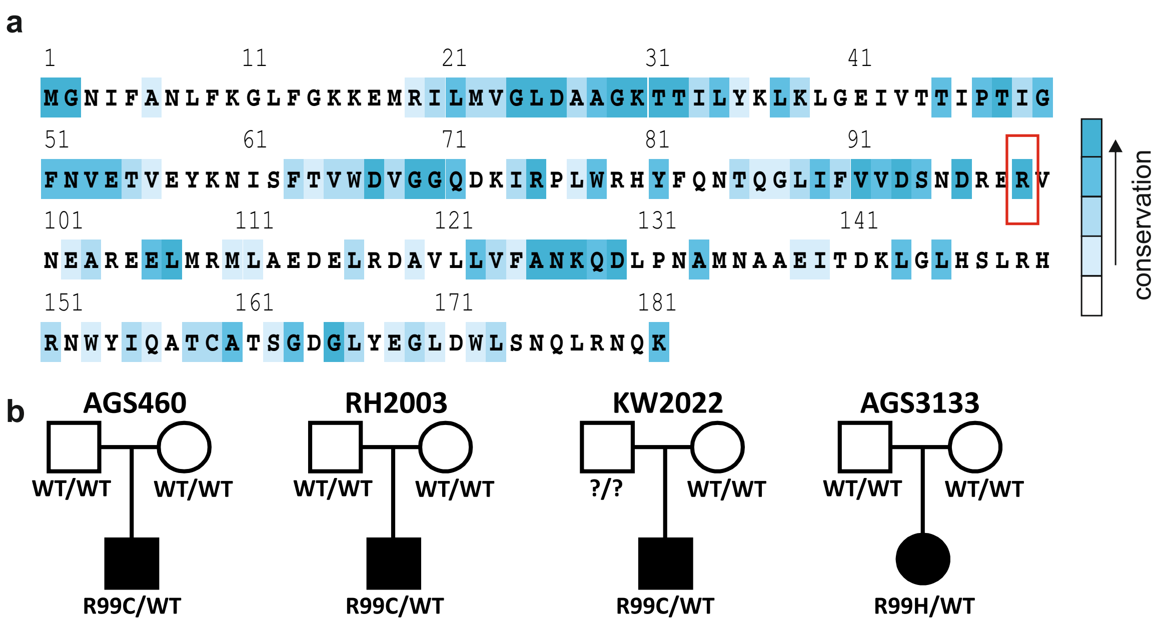

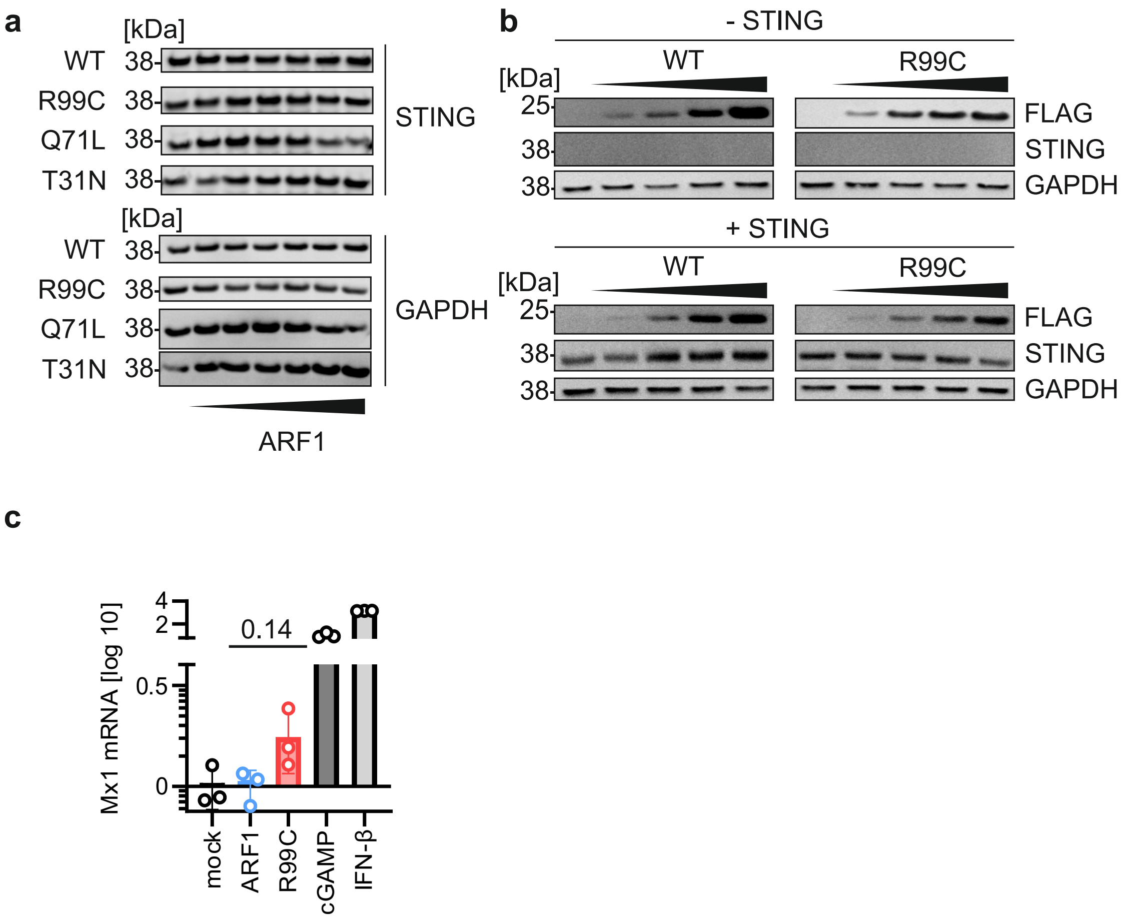
a
[kDa]
WT
R99C
Q71L
T31N
38
STING
GAPDH
ARF1
b
- STING
+ STING
WT
R99C
[kDa]
25
38
FLAG
STING
GAPDH
c
Mx1 mRNA [log 10]
4
2
0.5
0
0.14
mock
ARF1
R99C
cGAMP
IFN-β

### Extended Data Fig. 3

Hirschenberger et al.

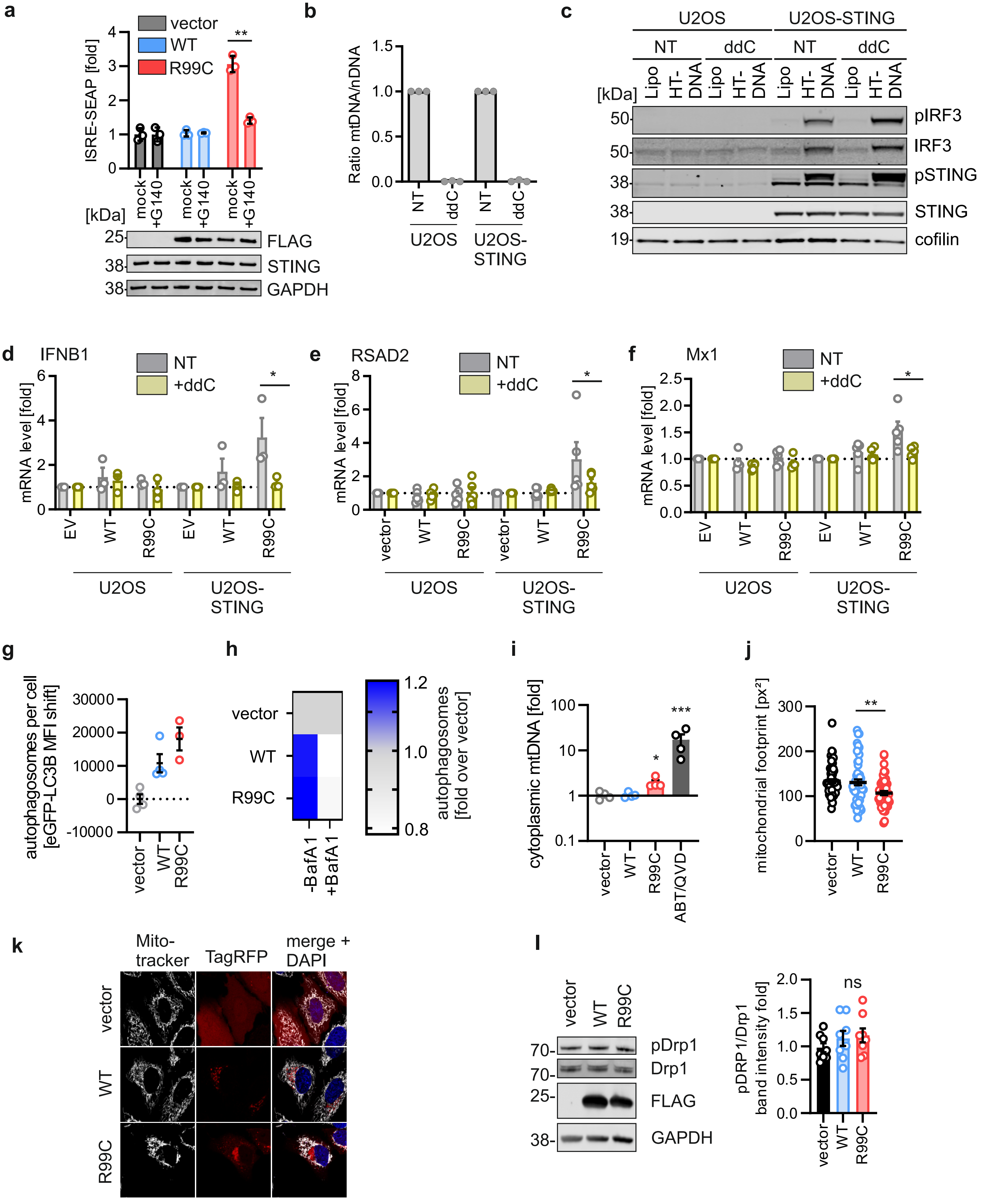

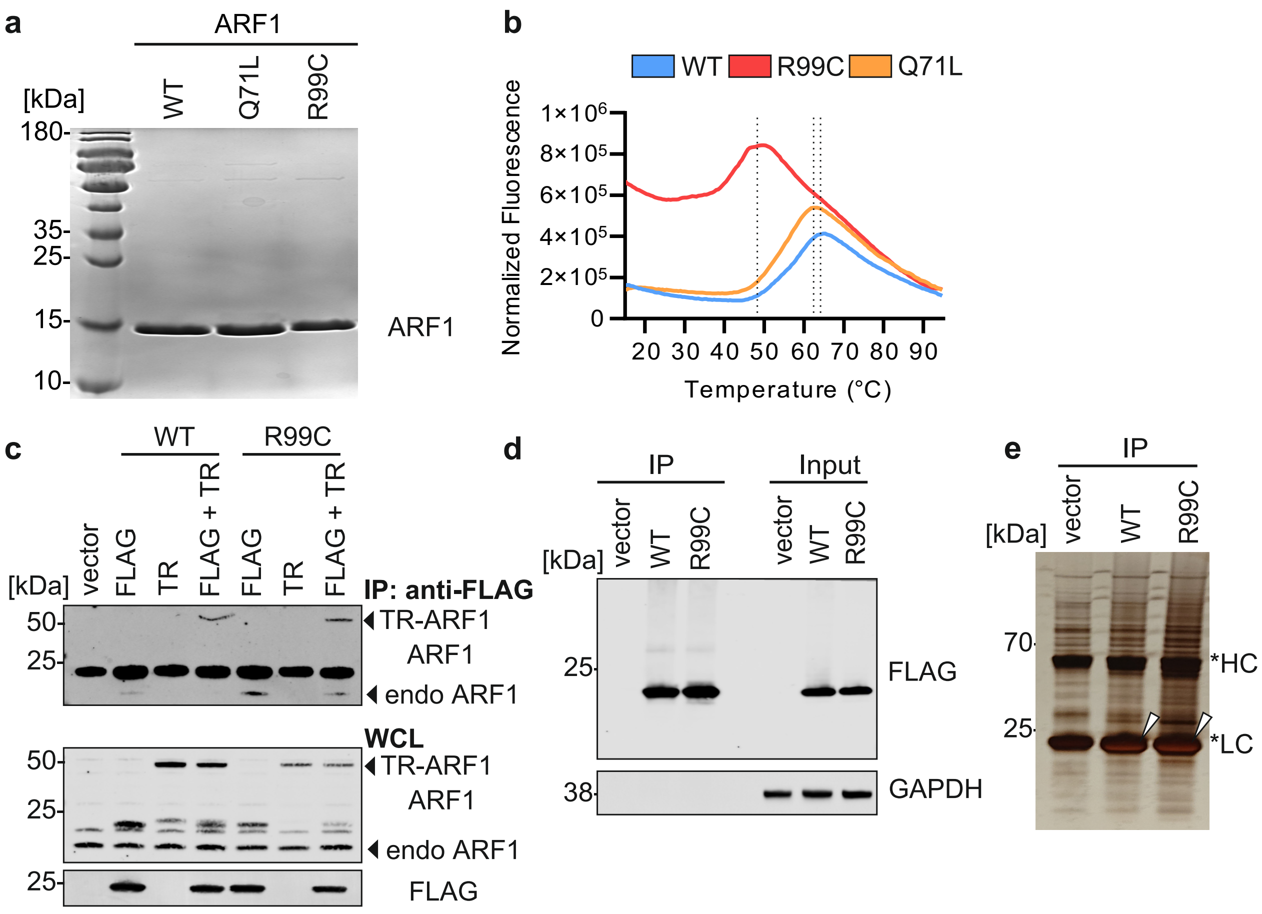
a
ARF1
WT
Q71L
R99C
[kDa]
180-
35-
25-
15-
10-
ARF1
b
WT
R99C
Q71L
Normalized Fluorescence
1×10^6
8×10^5
6×10^5
4×10^5
2×10^5
0
20 30 40 50 60 70 80 90
Temperature (°C)
c
WT
R99C
vector
FLAG
TR
FLAG + TR
FLAG
TR
FLAG + TR
[kDa]
IP: anti-FLAG
50-
25-
TR-ARF1
ARF1
endo ARF1
WCL
50-
25-
TR-ARF1
ARF1
endo ARF1
25-
FLAG
d
IP
Input
vector
WT
R99C
vector
WT
R99C
[kDa]
25
FLAG
38
GAPDH
e
IP
vector
WT
R99C
[kDa]
70
25
*HC
*LC

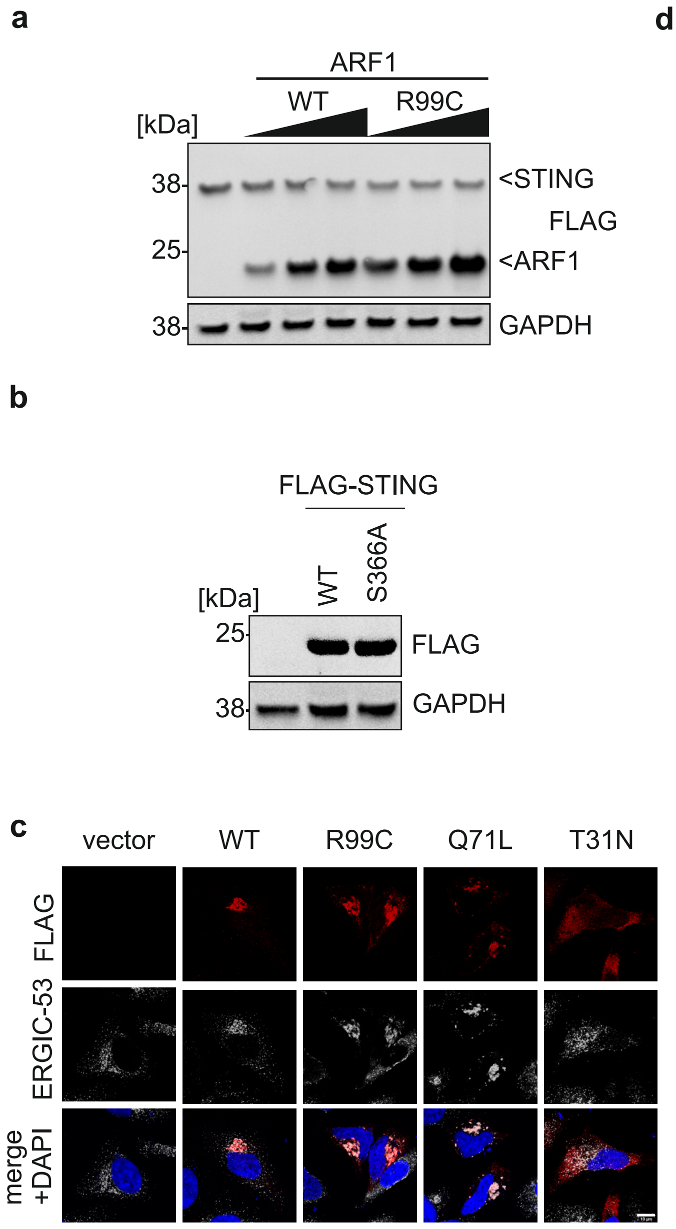
a
ARF1
WT
R99C
[kDa]
38-
25-
38-
<STING
FLAG
<ARF1
GAPDH
b
FLAG-STING
WT
S366A
[kDa]
25-
38-
FLAG
GAPDH
c
vector
WT
R99C
Q71L
T31N
FLAG
ERGIC-53
merge +DAPI

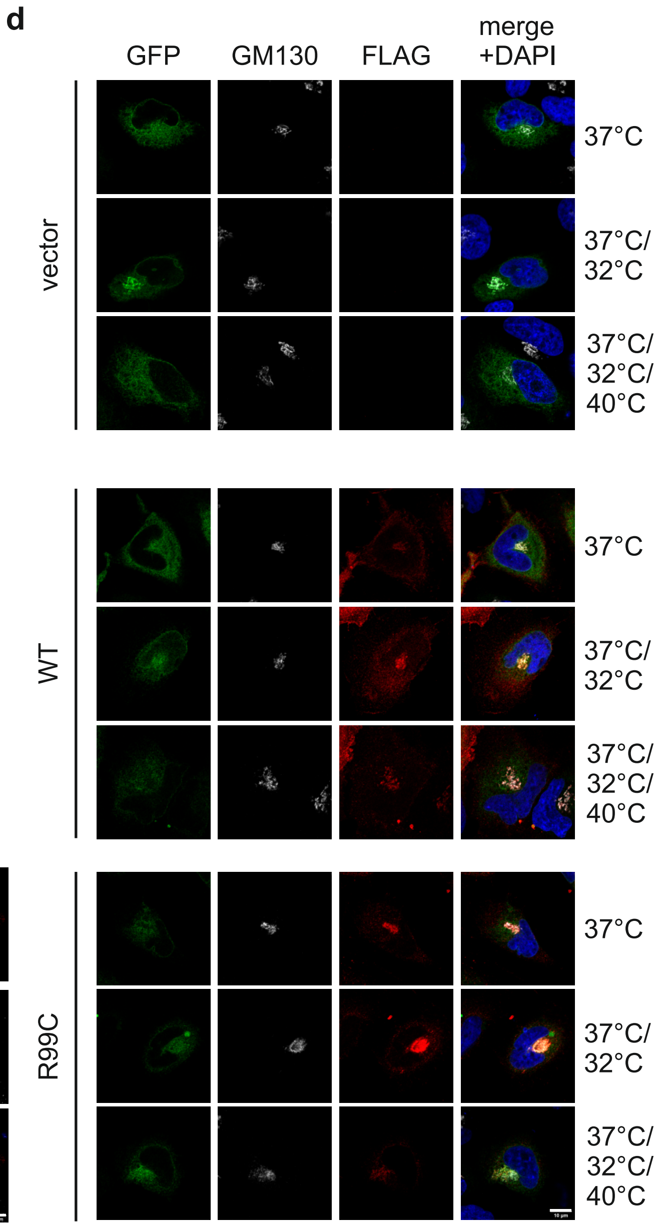
d
GFP
GM130
FLAG
merge +DAPI
vector
WT
R99C
37°C
37°C/ 32°C
37°C/ 32°C/ 40°C

### Extended Data Fig. 6

Hirschenberger et al.

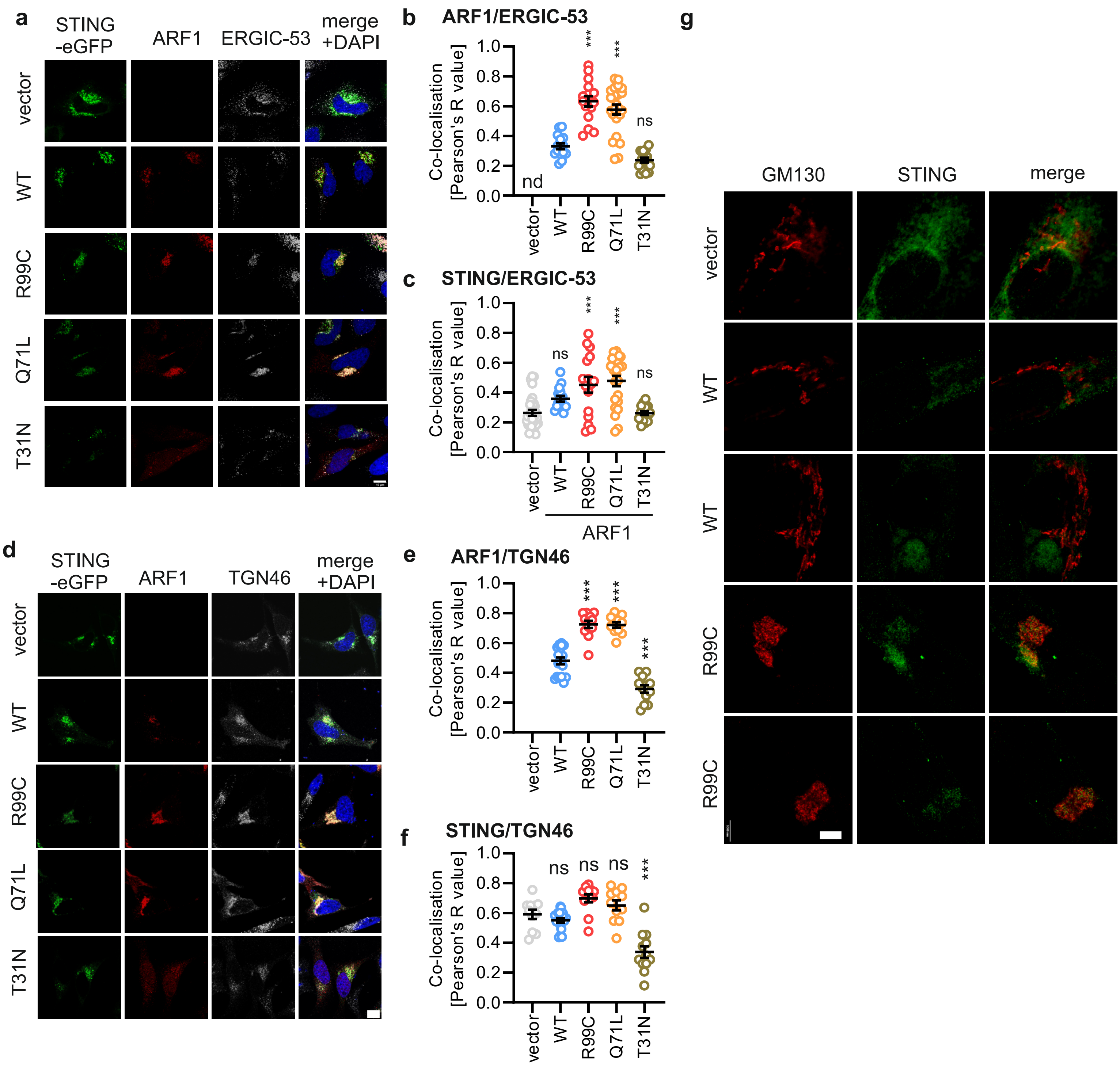

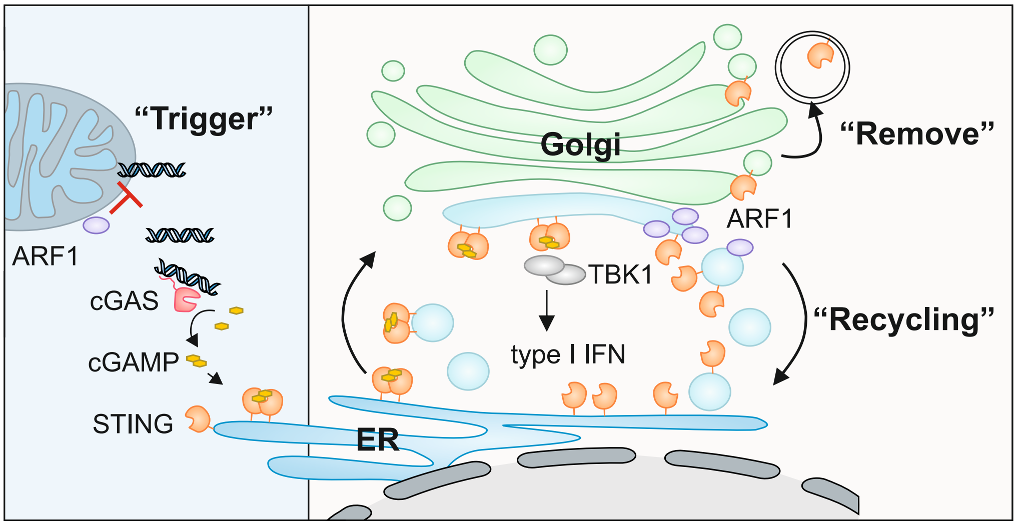

“Trigger”
ARF1
cGAS
cGAMP
STING
Golgi
“Remove”
ARF1
TBK1
type I IFN
“Recycling”
ER
